# Neurodevelopmental outcomes at one year in offspring of mothers who test positive for SARS-CoV-2 during pregnancy

**DOI:** 10.1101/2021.12.15.21267849

**Authors:** Andrea G. Edlow, Victor M. Castro, Lydia L. Shook, Anjali J. Kaimal, Roy H. Perlis

## Abstract

**Importance:** Epidemiologic studies suggest maternal immune activation during pregnancy may be associated with neurodevelopmental effects in offspring.

**Objective:** To determine whether in utero exposure to the novel coronavirus SARS-CoV-2 is associated with risk for neurodevelopmental disorders in the first 12 months after birth.

**Design:** Retrospective cohort

**Participants:** Live offspring of all mothers who delivered between March and September 2020 at one of six Massachusetts hospitals across two health systems.

**Exposure:** SARS-CoV-2 infection confirmed by PCR during pregnancy

**Main Outcome and Measures:** Neurodevelopmental disorders determined from ICD-10 diagnostic codes over 12 months; sociodemographic and clinical features of mothers and offspring; all drawn from the electronic health record.

**Results:** The cohort included 7,772 live births (7,466 pregnancies, 96% singleton, 222 births to SARS-CoV-2 positive mothers), with mean maternal age of 32.9 years; offspring were 9.9% Asian, 8.4% Black, and 69.0% white; 15.1% were of Hispanic ethnicity. Preterm delivery was more likely among exposed mothers (14% versus 8.7%; p=.003). Maternal SARS-CoV-2 positivity during pregnancy was associated with greater rate of neurodevelopmental diagnoses (crude OR 2.17 [95% CI 1.24-3.79, p=0.006]) as well as models adjusted for race, ethnicity, insurance status, offspring sex, maternal age, and preterm status (adjusted OR 1.86 [95% CI 1.03-3.36, p=0.04]). Third-trimester infection was associated with effects of larger magnitude (adjusted OR 2.34, 95% CI 1.23-4.44, p=0.01)

**Conclusion and Relevance:** Our results provide preliminary evidence that maternal SARS-CoV-2 may be associated with neurodevelopmental sequelae in some offspring. Prospective studies with longer follow-up duration will be required to exclude confounding and confirm these effects.

**Trial Registration:** NA

**Key Points:** *Question:* Does COVID-19 exposure in utero increase the risk for neurodevelopmental disorders in the first year of life?

*Findings:* In a cohort of babies delivered during COVID-19, those born to mothers with a positive SARS-CoV-2 PCR test during pregnancy were more likely to receive a neurodevelopmental diagnosis in the first 12 months after delivery, even after accounting for preterm delivery.

*Meaning:* These preliminary findings suggest that COVID-19 exposure may impact neurodevelopment, and highlight the need for prospective investigation of outcomes in children exposed to COVID-19 in utero.

## Introduction

The potential impact of maternal COVID-19 infection on offspring, if any, is not yet understood. However, the profound immune activation observed in a subset of infected individuals suggests that, even if the virus itself rarely crosses the placenta, the developing fetal brain may be impacted by maternal and placental inflammation and altered cytokine expression during key developmental windows ^1-3^. Epidemiologic studies demonstrate that maternal infection in pregnancy, including viral infections such as influenza, is associated with adverse neurodevelopmental outcomes in offspring, including autism spectrum disorders, schizophrenia, cerebral palsy, cognitive dysfunction, bipolar disorder, and anxiety and depression^4-9^. While the magnitude of these effects and strength of association varies, the consistency of such associations is difficult to ignore.

As some of these disorders may not manifest until adolescence or adulthood, the true risks of maternal immune activation may not become apparent for decades. However, convergent evidence suggests reason for concern. For example, two small brain imaging studies in children using either [18F]-FDG-PET^1^ or MRI^2^ suggested abnormalities following acute infection with SARS-CoV-2. More broadly, in both adults^3456^ and children^7^, a subset of individuals manifest neuropsychiatric symptoms after COVID-19 that can persist up to a year after acute illness. To begin to understand the neurodevelopmental impact of maternal SARS-CoV-2 infection, we examined electronic health records to provide preliminary estimates of risk, comparing offspring of SARS-CoV-2-infected mothers to offspring of those without, accounting for other potential confounding features.

## Methods

### Study design and data set generation

We extracted data from the electronic health records (EHR) of two academic medical centers and six community hospitals: Massachusetts General Hospital (MGH), Brigham and Women’s Hospital (BWH), Newton-Wellesley Hospital (NWH), North Shore Medical Center (NSMC), Martha’s Vineyard Hospital (MVH), Nantucket Cottage Hospital (NCH), Cooley Dickinson Hospital (CDH), and Wentworth Douglass Hospital (WDH) to identify all live births occurring between March and September 2020. Offspring were linked to maternal health records using data from the Electronic Data Warehouse (EDW) based upon date/time of birth, medical record number, and offspring sex. For mothers, we queried ICD-10 billing codes, problem lists, medications, and laboratory studies occurring from date of estimated last menstrual period up to the discharge date of the delivery admission, as well as sociodemographic features (maternal age, self-reported gender, insurance type, self-reported race and ethnicity based on US Census categories). Race and ethnicity were characterized to allow better control of confounding, recognizing that COVID-19 has differentially impacted these groups. For offspring, we also queried ICD-10 billing codes and problem lists. Data were managed with i2b2 server software (i2b2 v1.6.04, Boston, MA, USA)^12 -14^. The Mass General-Brigham Institutional Review Board approved all aspects of this study, with a waiver of informed consent as no patient contact was required, the study was considered to be minimal risk, and consent could not feasibly be obtained.

### Outcome definition

The primary outcome of interest was diagnosis of a neurodevelopmental disorder, based on presence of at least one ICD10 code included in the Healthcare Cost and Utilization Project (HCUP) level 2 ‘developmental’ category (code 654), including F8x (pervasive and specific developmental disorders: developmental disorders of speech and language [F80]; specific developmental disorders of scholastic skills [F81]; specific developmental disorder of motor function [F82]; pervasive developmental disorders [F84]; other/unspecific disorder of psychological development [F88/89]) and F7x (intellectual disabilities). Charts for all positive cases among exposed offspring were reviewed independently by two physicians (RP, AE) to confirm documentation of corresponding diagnosis. Controls were defined as the absence of any of these codes. To further explore potential for confounding, we secondarily examined HCUP Level 2 categories not reflecting delivery complications or congenital anomalies with prevalence of at least 2% in exposed or unexposed pregnancies.

### Exposure definition

Maternal SARS-CoV-2 positivity was defined on the basis of laboratory PCR result at any point during pregnancy, at any of the hospital network laboratories or tests at outside laboratories imported into the EMR. (During the period under investigation, routine testing of asymptomatic pregnant women was not conducted). Exploratory analysis examined trimester of exposure. Exposure trimester was estimated based on the established gestational age in the EMR: first trimester (0-12 weeks’ gestation) second trimester (12-26 weeks) and third trimester (26 weeks to delivery). Those individuals with no documented positive PCR results were considered to be negative.

### Analysis

We fit logistic regression models associating maternal SARS-CoV-2 status with the neurodevelopmental outcome, then added maternal age in years, race and ethnicity, insurance type (public versus private), as well as offspring sex and preterm status, to yield unadjusted and adjusted estimates of effect and 95% confidence intervals. To account for multiple births, they were considered to be clustered within deliveries; we used glm.cluster in the R *miceadds* package (v3.11-6) to generate robust standard errors. Sensitivity analyses restricting the cohort to full-term deliveries, or estimating effects limited to exposure in third trimester (excluding offspring with exposures in the other trimesters), used the same analytic approach. Additional sensitivity analyses to detect confounding examining other 12-month outcomes likewise applied crude and adjusted models with the same covariates.

All analyses utilized R 4.0.3 (The R Foundation for Statistical Computing, Vienna, Austria). Statistical significance for the primary outcome was defined as uncorrected two-tailed p<0.05; results for exploratory analyses apply the same threshold. As an EHR study, no observations were missing. E-value^8^ – the magnitude of association between a confounder and the exposure, and the confounder and outcome, required to yield the observed association if the true association is 1 – was calculated using *Evalue* 4.1.3 in R^9^.

## Results

Characteristics of the SARS-CoV-2 exposed mother-offspring pairs, as well as the unexposed pairs, are summarized in Table 1. The cohort included 7,772 live births (7,466 pregnancies, 96% singleton), with mean maternal age of 32.9 years; they were 9.9% Asian, 8.4% Black, and 69.0% white; 15.1% were of Hispanic ethnicity. The overall rate of SARS-CoV-2 positivity in pregnancy was 2.9% (222/7772). Zero exposed and one unexposed offspring were deceased before 12 months, and were excluded from analysis. Exposed mothers were significantly less likely to be of white or Asian race, more likely to be of Hispanic ethnicity, and more likely to have public versus private insurance. Rates of diabetes and hypertension were similar between the two groups. Preterm delivery was significantly more likely among exposed mothers (14% versus 8.7%; p=.003).

**Table 1.** Sociodemographic and clinical characteristics of maternal and offspring study groups

| Characteristic | Pregnancy SARS-COV-2 Negative, N = 7,550 | Pregnancy SARS-COV-2 Positive, N = 222 | p-value<sup>1</sup> |
| --- | --- | --- | --- |
| **Maternal age, Median (IQR)** | 33.0 (30.0 – 36.0) | 31.0 (26.2 – 35.0) | <0.001 |
| **Maternal race, n (%)** |  |  | <0.001 |
| Asian | 765 (10) | 7 (3.2) |  |
| Black or African American | 617 (8.2) | 39 (18) |  |
| Other | 657 (8.7) | 76 (34) |  |
| Unknown | 230 (3.0) | 18 (8.1) |  |
| White | 5,281 (70) | 82 (37) |  |
| **Maternal ethnicity, n (%)** |  |  | <0.001 |
| Hispanic | 1,019 (13) | 115 (52) |  |
| Not Hispanic | 6,283 (83) | 95 (43) |  |
| Unavailable | 248 (3.3) | 12 (5.4) |  |
| **Maternal public insurance, n (%)** | 1,297 (17) | 134 (60) | <0.001 |
| **Trimester of maternal SARS-COV-2 infection, n (%)** |  |  | NA |
| 1st | 0 (NA) | 1 (0.5) |  |
| 2nd | 0 (NA) | 61 (27) |  |
| 3rd | 0 (NA) | 160 (72) |  |
| Unknown | 7,550 | 0 |  |
| **Maternal gestational diabetes, n (%)** | 1,025 (14) | 38 (17) | 0.13 |
| **Maternal pre-eclampsia, n (%)** | 1,398 (19) | 43 (19) | 0.75 |
| **Maternal hemorrhage, n (%)** | 1,512 (20) | 41 (18) | 0.57 |
| **Delivery hospital type, n (%)** |  |  | <0.001 |
| Academic medical center | 4,383 (58) | 165 (74) |  |
| Community hospital | 3,167 (42) | 57 (26) |  |
| **Delivery method, n (%)** |  |  | 0.86 |
| C-Section | 2,423 (32) | 70 (32) |  |
| Vaginal | 5,127 (68) | 152 (68) |  |
| **Multiple births, n (%)** | 292 (3.9) | 14 (6.3) | 0.066 |
| **Delivery admission length of stay (days), Median (IQR)** | 3.00 (2.00 – 3.00) | 3.00 (2.00 – 3.00) | 0.85 |
| **Pre-term birth, n (%)** |  |  | 0.003 |
| Preterm | 654 (8.7) | 32 (14) |  |
| Term | 6,896 (91) | 190 (86) |  |
| **Offspring gender, n (%)** |  |  | 0.58 |
| Female | 3,714 (49) | 105 (47) |  |
| Male | 3,836 (51) | 117 (53) |  |
| **Offspring gestational age (weeks), Median (IQR)** | 39.29 (38.43 – 40.14) | 39.14 (37.71 – 40.11) | 0.013 |
| Unknown | 2 | 0 |  |
| **Offspring birth weight (grams), Median (IQR)** | 3,345 (3,015 – 3,655) | 3,257 (2,895 – 3,544) | 0.001 |
| Unknown | 55 | 4 |  |
| **Offspring birth length (inches), Median (IQR)** | 20.00 (19.00 – 20.50) | 19.50 (19.00 – 20.08) | 0.015 |
| Unknown | 244 | 15 |  |
| **Offspring 1-min. APGAR score, Median (IQR)** | 8.00 (8.00 – 9.00) | 8.00 (8.00 – 8.00) | 0.052 |
| **Offspring 5-min. APGAR score, Median (IQR)** | 9.00 (9.00 – 9.00) | 9.00 (9.00 – 9.00) | 0.85 |

<sup>1</sup>Wilcoxon rank sum test; Pearson's Chi-squared test; Fisher's exact test

In all, 14/222 exposed offspring (6.3%), and 227/7,550 unexposed offspring (3.0%), received a neurodevelopmental diagnosis within 12 months; crude OR 2.17 [95% CI 1.24-3.79, p=0.006]. Table 2 lists the most commonly-observed neurodevelopmental diagnoses by case or control status, including specific developmental disorder of motor function (F82), expressive language disorder (F80.1), and developmental disorder of speech and language, unspecified (F80.9). Median time to diagnosis was about two months earlier among exposed compared to unexposed offspring: 214 days [IQR 133] vs 275 days [IQR 93].

**Table 2.**
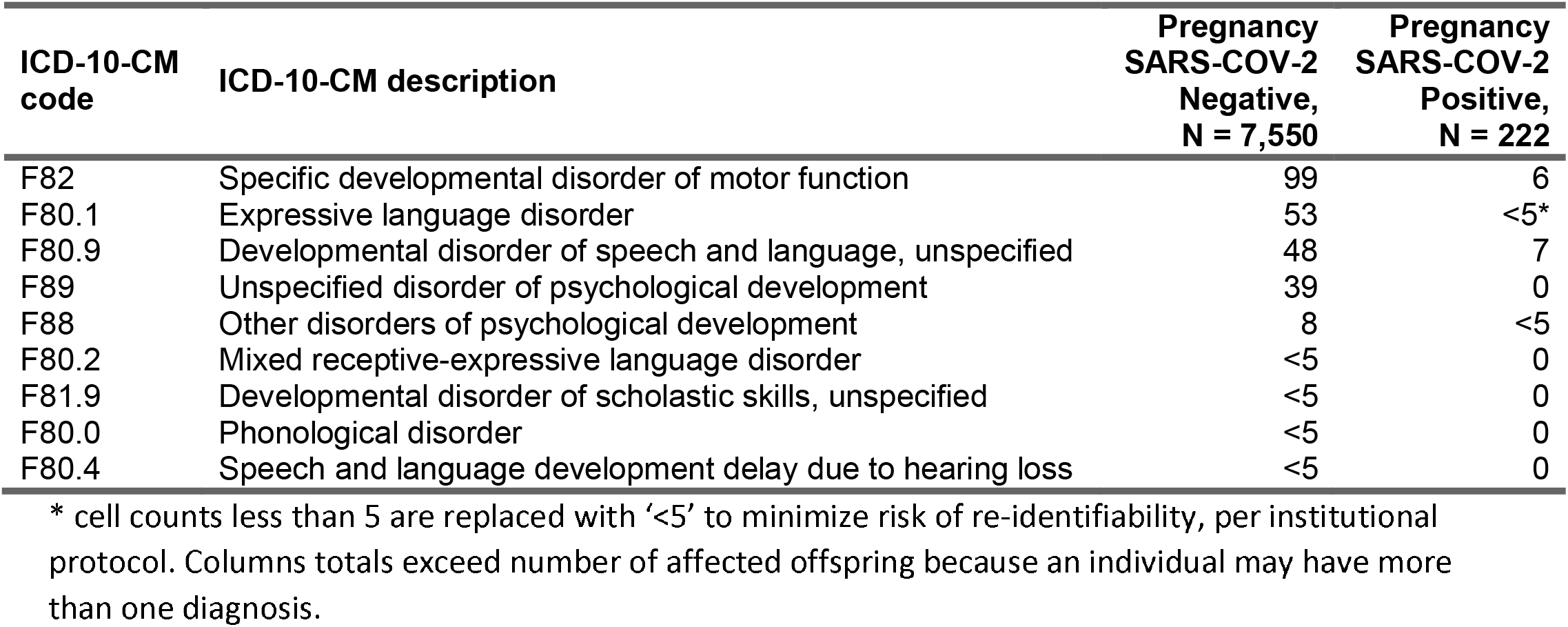
Frequency of individual developmental disorder ICD-10-CM codes in cases and controls.

In fully-adjusted regression models, accounting for non-singleton deliveries as clustered within-delivery, OR for any neurodevelopmental diagnosis among COVID-exposed offspring was 1.86 [95% CI 1.03-3.36, p=0.04]; Figure 1. In sensitivity analysis, we examined the contribution of preterm delivery to observed risk. Without adjusting for preterm delivery, but with all other covariates included, adjusted OR was 1.97 (95% CI 1.10-3.50; p=.024). When analysis was limited to full-term pregnancies (n=6896 unexposed, 190 exposed offspring) fully-adjusted OR was 1.68 (0.81-3.45; p=.16); Supplemental Figure 1. We also compared offspring of mothers infected in third trimester alone to those of uninfected mothers, with exclusion of offspring of mothers infected in first or second trimester, yielding adjusted OR of 2.34 (95% CI 1.23-4.44) (Supplemental Figure 2).

**Figure 1.**
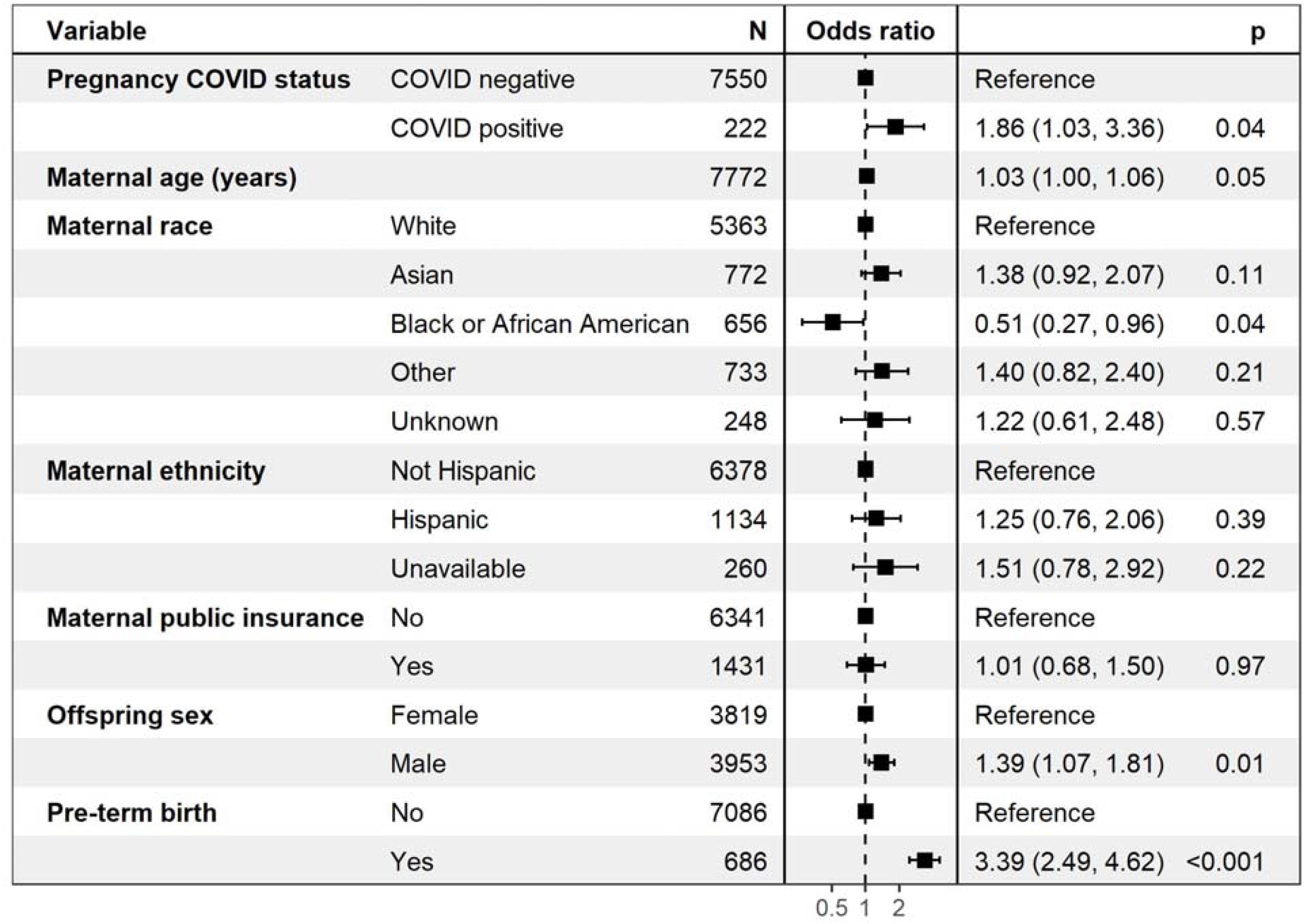
Forest plot of adjusted model for risk of offspring developmental disorder.

In addition to these sensitivity analyses, we sought to quantify the possibility that results reflected confounding by other aspects of maternal sociodemographic status or comorbidity not captured in our regression models. First, we calculated the E-value for the observed association^810^, yielding 3.12 (95% CI, 1.21-NA), indicating that an undetected confounder would need to be 3.12 times more common among the exposed, and cause a 3.12-fold increase in risk, to yield the observed effect if true OR was 1. We also examined rates of non-neurodevelopmental diagnostic categories with frequency of at least 2% in either the exposed or unexposed groups, again using unadjusted and then adjusted multiple logistic regression models (Supplemental Table 1), anticipating that associations would be inflated by an unmeasured confounder. In adjusted models, among 27 diagnostic categories, only viral infection was significantly more common among exposed offspring (crude OR 2.59 (95% CI 1.76-3.81), adjusted OR 1.81 (1.20-2.72).

## Discussion

In this analysis of 222 offspring of SARS-CoV-2 infected mothers, compared to the offspring of 7,550 of uninfected control mothers delivered during the same period, we observed neurodevelopmental diagnoses to be significantly more common among exposed offspring, particularly those exposed to 3^rd^ trimester maternal infection. The majority of these diagnoses reflected developmental disorders of motor function or speech and language. Notably, while we identify greater risk of preterm delivery among SARS-CoV-2 positive mothers as in prior studies^1112^, adjustment for preterm birth does not account for all of the observed increased risk of incurring a neurodevelopmental diagnosis – i.e., the fully-adjusted regression model including preterm delivery still indicates elevated risk, with adjusted OR of 1.86 for any neurodevelopmental diagnosis among COVID-exposed offspring. Moreover, the magnitude of this effect is only modestly diminished among infants delivered at ≥37 weeks, with AOR 1.68. Of note, given the known association between severe COVID-19 in pregnancy and increased risk for preterm birth, those excluded in this sensitivity analysis are theoretically the individuals most at risk for adverse neurodevelopmental programming based on the proposed mechanism. The finding that the directionality and magnitude of effect is maintained among term deliveries provides further evidence that this association requires follow-up in larger studies adequately powered for such an analysis.

Whether a definitive connection exists between prenatal SARS-CoV-2 exposure and adverse neurodevelopment in offspring is not yet known, in part because children born to women infected in the first wave of the pandemic are under 2 years old. A longitudinal cohort study of 57 infants with prenatal exposure to SARS-CoV-2 in China identified deficits in social-emotional domain of neurodevelopmental testing at 3 months of age, although the study design did not permit controlling for important confounders such as mother-baby separation nor did it include a non-infected comparator group^13^. The authors of a recent pre-print report of a prospective cohort study of 238 infants born during the pandemic (both exposed and non-exposed to SARS-CoV-2 during pregnancy) at 6 months argue that observed neurodevelopmental deficits in both groups may be the product of maternal pregnancy during the pandemic itself, rather than SARS-CoV-2 exposure per se^14^. The biological basis/mechanism by which maternal pandemic-associated stress would be a more dominant driver of offspring neurodevelopment than maternal viral illness in pregnancy remains unclear, and this putative association also requires validation in larger and longer-term studies. Our findings identifying an association between prenatal SARS-CoV-2 exposure and neurodevelopmental diagnoses at 12 months are consistent with a large body of literature including human and animal studies linking maternal viral infection and maternal immune activation with offspring neurodevelopmental disorders later in life^4 -9^, some of which can be foreshadowed as early as the first year of life^15^.

### Limitations

Our results must be recognized as preliminary given the limited duration of follow-up. In particular, we cannot exclude the possibility that additional neurodevelopmental effects will become apparent later in life – indeed, the offspring analyzed here are younger than the age at which neurodevelopmental disorders such as autism are typically diagnosed. Conversely, there may be a form of ascertainment bias arising from greater concern for offspring of mothers who were ill during pregnancy – that is, parents may be more inclined to seek evaluation, or clinicians more inclined to diagnose or refer for evaluation. Our retrospective study design and reliance on ICD-10 diagnosis codes also lacks the sensitivity of a prospective cohort study that incorporates detailed neurocognitive phenotyping; such studies will be important to better define the impact, if any, of maternal SARS-CoV-2 infection. As an ‘open’ health system, we cannot exclude the possibility of misclassification, as mothers classified as SARS-CoV-2 negative may have received a positive test result or care for SARS-CoV-2 illness outside of our system, and offspring may receive follow-up in another health system. Such misclassification should occur completely at random – i.e., there is no clear reason that SARS-CoV-2-exposed offspring delivered in a specific health system would be less likely to receive ongoing care in that system. In general, these effects of misclassified exposure or outcome would tend to bias our results toward the null hypothesis. We also note that our overall rate of SARS-CoV-2 positivity in pregnancy is lower than has been reported elsewhere^16^, likely reflecting the inclusion of smaller and community hospitals together with urban academic medical centers in our cohort.

### Conclusion

These preliminary findings suggest greater risk for adverse neurodevelopmental outcomes at one year among SARS-CoV-2 exposed offspring, and highlight the urgency of follow-up studies in large and representative cohorts. More broadly, our analysis indicates the feasibility of leveraging EHR data for an in silico cohort that may enable detection of risk signals before such large-scale, prospective follow-up studies are available. The approach described here, using coded clinical data extracted from the EHR, is amenable to scaling across multiple health systems in the US and internationally. Such follow-up studies will be critical in confirming the associations we identify, and more precisely estimating the risk for, and potential nature of, neurodevelopmental sequelae of in utero exposure to SARS-CoV-2.

## Data Availability

Data are not available for external release.

## Acknowledgements

This study was supported by the National Institute of Mental Health (R01MH116270 and 1R56MH115187; Dr. Perlis) and the National Institute of Child Health and Human Development (R01 HD100022-02S2; Dr. Edlow). The sponsors did not contribute to any aspect of the design and conduct of the study; collection, management, analysis, and interpretation of the data; preparation, review, or approval of the manuscript; and decision to submit the manuscript for publication. The authors had the final responsibility for the decision to submit for publication. Dr. Perlis had full access to all the data in the study and takes responsibility for the integrity of the data and the accuracy of the data analysis.

Dr. Perlis has received consulting fees from Burrage Capital, Genomind, RID Ventures, Belle Artificial Intelligence, and Takeda. He holds equity in Psy Therapeutics, Belle Artificial Intelligence, and Circular Genomics. The other authors report no disclosures.

## Author Contributions

Andrea G. Edlow, MD MSc – drafted manuscript, revised manuscript

Victor M. Castro, MS – conducted data cleaning and analysis, revised manuscript

Lydia L. Shook, MD – revised manuscript

Anjali J. Kaimal, MD MAS – planned the analysis, revised manuscript

Roy H. Perlis, MD MSc – planned the analysis, drafted manuscript, revised manuscript

### Data Availability Statement

Data not available.

