## Supplemental Materials for "Neurodevelopmental outcomes at one year in offspring of mothers who test positive for SARS-CoV-2 during pregnancy"

**Affiliations:**

1. Department of Obstetrics and Gynecology, Massachusetts General Hospital and Harvard Medical School, Boston, MA.

2. Center for Quantitative Health and Department of Psychiatry, Massachusetts General Hospital and Harvard Medical School, Boston, MA.

3. Research Information Science and Computing, Mass General Brigham, Somerville, MA.

**Correspondence:**

Roy H. Perlis

Department of Psychiatry

Massachusetts General Hospital

Simches Research Building

185 Cambridge St

Boston, MA 02114

rperlis@[mgh.harvard.edu](https://www.dropbox.com/referrer_cleansing_redirect?hmac=ZU1%2FOUFWNSQ7Q0U%2Fr10rbNg5n%2FmNthqiCpxLlJtZSQk%3D&url=http%3A%2F%2Fpartners.org)

(617) 726-7426

**Abstract**

**Importance:** Epidemiologic studies suggest maternal immune activation during pregnancy may be associated with neurodevelopmental effects in offspring.

**Objective:** To determine whether in utero exposure to the novel coronavirus SARS-CoV-2 is associated with risk for neurodevelopmental disorders in the first 12 months after birth.

**Design:** Retrospective cohort

**Participants:** Live offspring of all mothers who delivered between March and September 2020 at one of six Massachusetts hospitals across two health systems.

**Exposure:** SARS-CoV-2 infection confirmed by PCR during pregnancy

**Main Outcome and Measures:** Neurodevelopmental disorders determined from ICD-10 diagnostic codes over 12 months; sociodemographic and clinical features of mothers and offspring; all drawn from the electronic health record.

**Results:** The cohort included 7,772 live births (7,466 pregnancies, 96% singleton, 222 births to SARS-CoV-2 positive mothers), with mean maternal age of 32.9 years; offspring were 9.9% Asian, 8.4% Black, and 69.0% white; 15.1% were of Hispanic ethnicity. Preterm delivery was more likely among exposed mothers (14% versus 8.7%; p=.003). Maternal SARS-CoV-2 positivity during pregnancy was associated with greater rate of neurodevelopmental diagnoses (crude OR 2.17 [95% CI 1.24-3.79, p=0.006]) as well as models adjusted for race, ethnicity, insurance status, offspring sex, maternal age, and preterm status (adjusted OR 1.86 [95% CI 1.03-3.36, p=0.04]). Third-trimester infection was associated with effects of larger magnitude (adjusted OR 2.34, 95% CI 1.23-4.44, p=0.01)

**Conclusion and Relevance:** Our results provide preliminary evidence that maternal SARS-CoV-2 may be associated with neurodevelopmental sequelae in some offspring. Prospective studies with longer follow-up duration will be required to exclude confounding and confirm these effects.

**Figure S1.** Forest plot of adjusted model for risk of offspring developmental disorder excluding preterm deliveries*

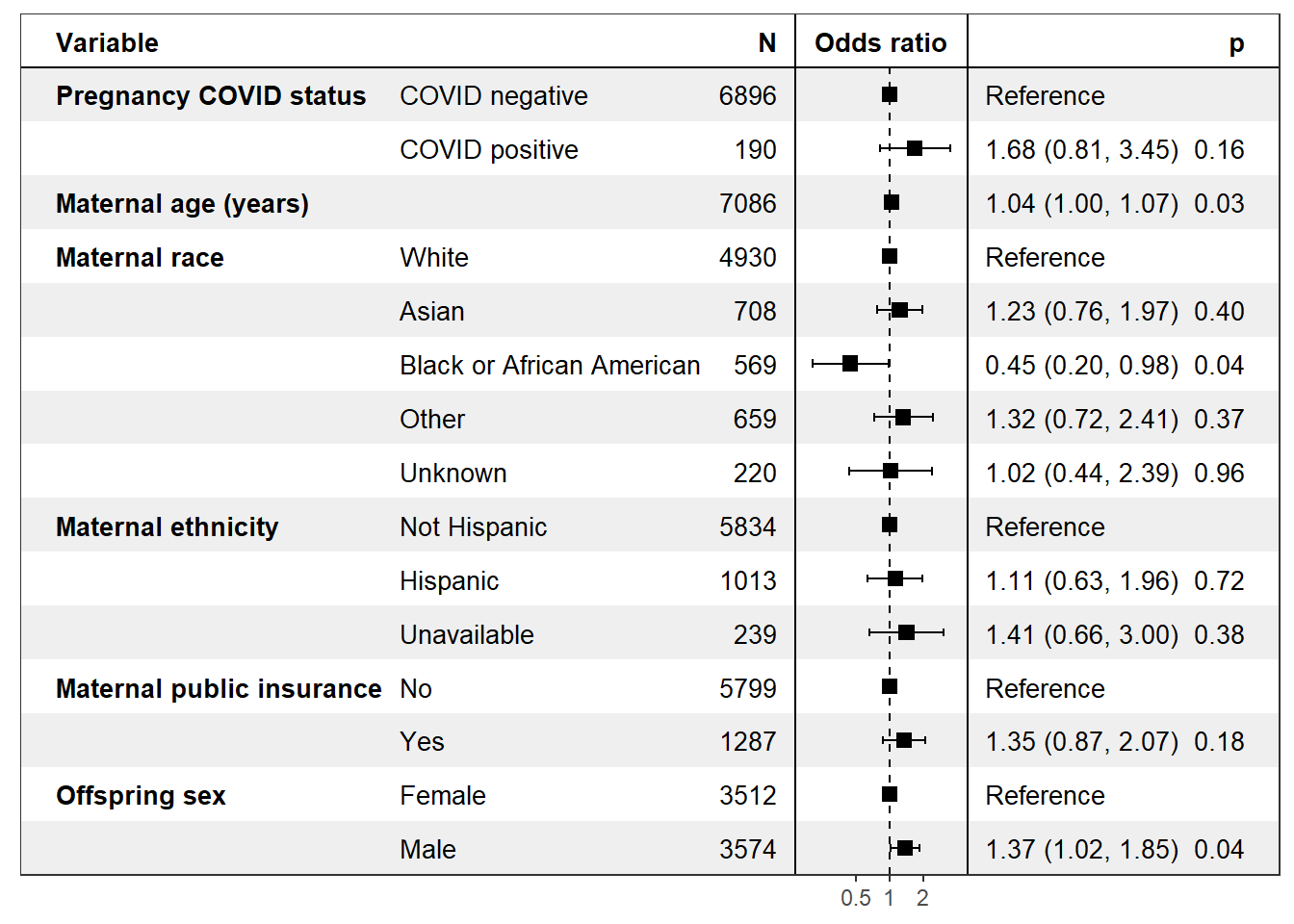

*****preterm deliveries defined as deliveries < 37 weeks’ gestation

**Figure S2.** Forest plot of adjusted model for risk of offspring developmental disorder excluding mothers infected in the first or second trimester (62 mothers excluded).

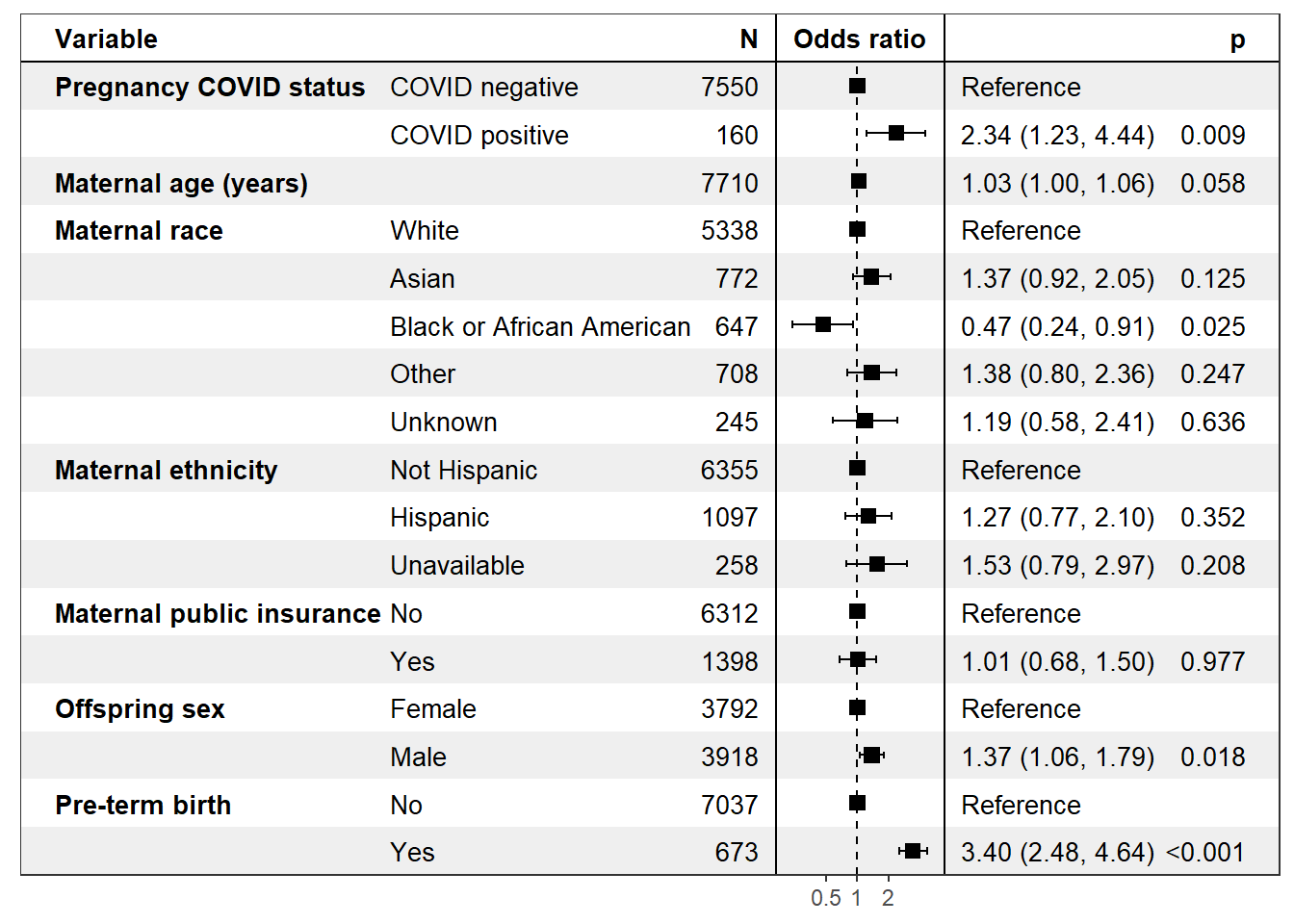

**Table S1.** Unadjusted and adjusted models for pregnancy SARS-COV-2 infection risk of offspring diagnosis (restricted to CCS diagnosis with greater than 3% in both cases and control group)

| **Diagnostic category** | **Pregnancy SARS-COV-2  pos,**  **N=222** | **Pregnancy SARS-COV-2  neg,**  **N=7,550** | **Unadj. OR (95% CI)** | **Unadj.  p-value** | **Adj. OR  (95% CI)** | **Adj. p-value** |
| --- | --- | --- | --- | --- | --- | --- |
| CCS 004 Mycoses | 14 | 283 | 1.73 (0.99-3.01) | 0.053 | 1.08 (0.60-1.91) | 0.805 |
| CCS 007 Viral infection | 32 | 461 | 2.59 (1.76-3.81) | <0.001 | 1.81 (1.20-2.72) | 0.005 |
| CCS 047 Other and unspecified benign neoplasm | 6 | 229 | 0.89 (0.39-2.02) | 0.777 | 0.96 (0.41-2.22) | 0.915 |
| CCS 051 Other endocrine disorders | <5 | 181 | 0.56 (0.18-1.76) | 0.319 | 0.53 (0.16-1.70) | 0.285 |
| CCS 058 Other nutritional; endocrine; and metabolic disorders | 35 | 805 | 1.57 (1.09-2.27) | 0.017 | 1.45 (0.99-2.13) | 0.059 |
| CCS 091 Other eye disorders | 13 | 329 | 1.37 (0.77-2.42) | 0.285 | 1.22 (0.68-2.21) | 0.505 |
| CCS 092 Otitis media and related conditions | 21 | 473 | 1.56 (0.99-2.47) | 0.056 | 1.51 (0.94-2.44) | 0.091 |
| CCS 094 Other ear and sense organ disorders | 14 | 278 | 1.76 (1.01-3.06) | 0.045 | 1.41 (0.79-2.52) | 0.241 |
| CCS 095 Other nervous system disorders | 9 | 194 | 1.60 (0.81-3.17) | 0.176 | 1.34 (0.66-2.72) | 0.417 |
| CCS 096 Heart valve disorders | 10 | 463 | 0.72 (0.38-1.37) | 0.319 | 0.64 (0.33-1.23) | 0.179 |
| CCS 117 Other circulatory disease | 8 | 214 | 1.28 (0.62-2.63) | 0.499 | 1.53 (0.73-3.22) | 0.261 |
| CCS 125 Acute bronchitis | 6 | 135 | 1.53 (0.67-3.49) | 0.318 | 1.34 (0.57-3.18) | 0.503 |
| CCS 126 Other upper respiratory infections | 29 | 688 | 1.50 (1.01-2.23) | 0.046 | 1.29 (0.85-1.95) | 0.236 |
| CCS 133 Other lower respiratory disease | 20 | 355 | 2.01 (1.25-3.22) | 0.004 | 1.57 (0.95-2.59) | 0.077 |
| CCS 134 Other upper respiratory disease | 12 | 483 | 0.84 (0.46-1.51) | 0.551 | 0.79 (0.43-1.45) | 0.452 |
| CCS 136 Disorders of teeth and jaw | 5 | 189 | 0.90 (0.37-2.20) | 0.813 | 0.78 (0.31-1.97) | 0.604 |
| CCS 138 Esophageal disorders | 9 | 323 | 0.95 (0.48-1.86) | 0.871 | 0.93 (0.47-1.87) | 0.848 |
| CCS 143 Abdominal hernia | 9 | 141 | 2.22 (1.12-4.42) | 0.023 | 1.30 (0.63-2.69) | 0.472 |
| CCS 151 Other liver diseases | 7 | 193 | 1.24 (0.58-2.67) | 0.581 | 1.11 (0.50-2.46) | 0.801 |
| CCS 155 Other gastrointestinal disorders | 40 | 816 | 1.81 (1.28-2.57) | <0.001 | 1.28 (0.88-1.85) | 0.194 |
| CCS 161 Other diseases of kidney and ureters | 5 | 120 | 1.43 (0.58-3.53) | 0.441 | 1.31 (0.51-3.35) | 0.579 |
| CCS 166 Other male genital disorders | 9 | 364 | 0.83 (0.42-1.64) | 0.599 | 1.05 (0.51-2.13) | 0.899 |
| CCS 198 Other inflammatory condition of skin | 9 | 258 | 1.19 (0.61-2.35) | 0.608 | 1.00 (0.50-2.01) | 0.996 |
| CCS 200 Other skin disorders | 31 | 753 | 1.47 (0.99-2.16) | 0.053 | 1.16 (0.77-1.73) | 0.480 |
| CCS 211 Other connective tissue disease | 8 | 151 | 1.83 (0.89-3.78) | 0.101 | 1.36 (0.64-2.89) | 0.424 |
| CCS 244 Other injuries and conditions due to external causes | 11 | 302 | 1.25 (0.68-2.32) | 0.477 | 1.21 (0.64-2.30) | 0.553 |
| CCS 654 Developmental disorders | 14 | 227 | 2.17 (1.24-3.79) | 0.006 | 1.86 (1.03-3.36) | 0.040 |
